# Gut microbial dysbiosis is associated with occurrence and post-surgical prognosis of unraptured cerebral aneurysm

**DOI:** 10.1101/2024.09.29.24314585

**Authors:** Jiancheng Du, Gang Li, Donghai Wang, Peng Zhao, Ping Zhang, Tianyi Dong, Junhao Wei, Lin Deng, Zhiping Dun, Shili Liu

## Abstract

**BACKGROUND:** The microbiota has been reported to play an important role in the occurrence of brain aneurysms (CA). However, no microbiota or metabolite has been used for diagnosis or therapy of CA till now. Therefore, we investigated the shifts of gut microbiota and metabolites in CA patients, and explored the feasibility of using them as biomarkers and therapeutic targets.

**METHODS:** The microbial DNA was extracted from the stool samples of the CA and healthy volunteer groups. The variable V3-V4 region of the 16S rDNA gene was sequenced on a MiSeq system. R software package was used to analyze the sequence, and the variations in microbial composition was obtained. The correlation analysis of the differential intestinal bacteria, metabolites and blood parameters was explored based on a logistic regression model. A random forest algorithm was used to predict the classification of samples for further exploring the relationship between fecal microbiota and CA.

**RESULTS:** The α-diversity indexes demonstrated an altered within-sample microbial diversities between patients and healthy people. The subsequent beta diversity results indicated that shift in the between-sample microbial diversities of the intestinal microbiota was associated with the occurrence of cerebral aneurysm. Specifically, the abundance of some gut bacterial genera, such as Blautia and Faecalibacterium, changed significantly after CA. Intestinal metabolite enrichment results highlight the role of carbohydrate metabolic pathways in CA, including sedoheptulose 7-phosphate (S-7-P). The correlation analysis of the differential intestinal bacteria, metabolites and blood parameters indicated that intestinal bacteria and metabolites were related to host blood parameters. Then, the predictive models were employed to test the significance of the combination of differential bacteria, metabolites and blood parameters in CA diagnosis and prognosis. The predictive model built by Faecalibacterium and s7p obtained an area under ROC curve (AUC) of 99.7%, and Faecalibacterium and s-7-p were also associated with some metabolite that have significance in post-surgery prognosis of CA.

**Conclusions:** The gut microbiota and metabolites profiles of patients with CA were significantly altered. Bacterial genus Faecalibacterium and metabolite s-7-p could serve as potential biomarkers in CA prediction and post-surgery prognosis.

## Background

Cerebral aneurysm (CA) are acquired, focal, saccular outpouchings of the artery wall, usually found at anterior communicating artery, posterior communicating artery, middle cerebral artery, anterior cerebral artery, vertebral artery, basilar artery, etc., and their overall prevalence in the general population is about 3.2% worldwide [1]. Among them, the incidences of unruptured CA in adults range from 1% to 8%. Once CA ruptured and bleeding, such as subarachnoid hemorrhage (SAH), etc., it will cause a serious cerebrovascular disease with extremely high disability and mortality rates. In China, the prevalence of unruptured CA is about 8.61% [2]. Statistics show that about 50% of patients with ruptured CA have poor prognosis, which can lead to death or serious disability sequelae [3]. According to the shape, CA is usually divided into two categories: saccular aneurysms and dissecting aneurysms.

Although more and more unruptured CA have been diagnosed around the world, current prevention and treatment measures are extremely limited due to the precise mechanism leading to the onset of CA remains unclear. Chronic inflammation was reported playing a crucial role in CA formation and rupture, several human CA pathology studies have demonstrated the presence of inflammatory cells and mediators in the CA wall and the subsequent deteriorated structure[4, 5]. Chronic inflammation is a multifactorial process involving both environmental and genetic factors. Studies of genetic susceptibility to cerebral aneurysm have identified that heritability, such as genes CDKN2A, SOX17, and ADAMTS15, play a moderate role [6, 7], suggesting that environmental factors also play an important role in the occurrence of CA.

There is growing evidence exhibits that gut microbiota is a key environmental factor affecting host metabolism and immune homeostasis. Recently, people’s attention is attracted to the role of the human microbiome in cardiovascular diseases such as hypertension, heart failure, and atherosclerosis[8-10]. Studies have shown that gut microbiota produces large amounts of metabolites in these diseases and are absorbed into the systemic circulation, where they are further metabolized by host enzymes, triggering damage to the target organ[11, 12]. Another recent investigation reported that depleting the gut microbiota using antibiotics reduced the incidence of CAs in mice [13]. The findings of a recent article suggested that *Hungatella hathewayi* associated taurine deficiency is a potential key factor in the pathogenesis of CA [14]. However, the role and exact mechanism of intestinal microbiota composition and metabolite variations in patients with CA remain inconclusive. As a result, there are no microbiota or metabolite biomarkers available for CA detection presently, as well no microbiological strategies to intervene against CA development.

As we know, there are many drawbacks existed in the present detection techniques of CA, such as CTA or MRA, which need the patient to inject contrast agents or require large instruments. In contrast, the human microbiota is a relatively easy to obtain sample, and their changes are closely related to the pathophysiological state of the body [15]. Certain microbial variations in microbiota can produce detectable changes earlier than the appearance of disease symptoms, which is an ideal biomarker for disease risk prediction. Exploring the correlation between microbiota and disease can contribute to providing insights for the interrogation of disease mechanism [16]. In clinical practice, after the diagnosis of unruptured CA, aneurysm clipping or interventional surgery is generally preferred, personalized comprehensive consideration is often made to guide clinical decision-making based on factors such as patient age, aneurysm size, morphology, location and type of aneurysm. Unfortunately, there is currently even no definitive biomarker for clipping or intervention as well as post-surgery prognosis for unruptured CA. Therefore, some patients at low risk of rupture may mistakenly assume the complications of surgical treatment, while some high-risk patients may develop aneurysm rupture due to failure to undergo surgery. If intestinal microbiota and its metabolites or blood parameters are found to be able to clearly indicate the formation of CA or post-surgery prognosis, this will be conducive to the prevention and subsequent treatment of CA, and the correlation within these parameters or between them and CA can also concomitantly provide clues for our in-depth understanding of CA formation.

Therefore, 108 patients diagnosed with unruptured CA and 40 healthy people were enrolled in the present study as research subjects, their blood and stool samples were collected. Correlations between the significantly altered ingredients in blood, fecal bacteria and metabolites were analyzed, as well the differential parameters were screened and used to construct prediction models, which were validated succeedingly with the new collected CA samples.

## Methods

### Patient samples collection and cell culture

To characterize the intestinal microbiota of the CA patients and healthy people, 108 patients with CA and 40 healthy volunteers were selected [40 controls from routine medical examination and 108 CA patients hospitalized in Qilu hospital, Shandong University (Jinan, Shandong, China) from February to October 2023; the CA patients consist 85 saccular aneurysms patients (A) and 23 dissecting aneurysms patients (D)]. All individuals recruited in the CA patient group were identified by Digital Subtraction Angiography (DSA) (**Supplemental Figure 1**), the recruited person’s basic information is exhibited in **Table 1**. Fecal samples were obtained from the volunteers, and microbial DNA was extracted from the stool samples of the two groups. The fecal samples were kept at − 80 °C for subsequent DNA extraction and sequencing.

### 16S sequencing

Total fecal genome DNA was extracted with CTAB/SDS method, concentration and purity of the extracted genome DNA was checked by agarose gels electrophoresis. DNA was diluted with sterile water to the concentration of 1 ng/μl. Using about 10 ng of this template DNA and 0.2 μM of primers that had adapters and barcodes, the variable V3-V4 region of the 16S rRNA gene was amplified by PCR (25 PCR cycles) with Phusion® High-Fidelity PCR Master Mix (New England Biolabs). Subsequently, amplicons were sequenced on a MiSeq system (Illumina, San Diego, CA). According to the unique barcodes, the Paired-end reads were assigned to each sample, and FLASH was used to merge the Paired-end reads. R software package (Quantitative Insights into Microbial Ecology) was employed for analysis of the sequences, and alpha- (within samples) and beta- (among samples) diversity were produced by in-house Perl scripts. In detail, reads were quality filtered by QIIME, and pick_de_novo_otus.py was used to make operational taxonomic units (OTUs) table. Sequences with ≥97% similarity were classified to an OTU. Then the representative sequence of an OTU was picked and taxonomic information was annotated using the RDP classifier. To obtain Alpha Diversity, the OTU table was rarified into three metrics: Chao1 represents the species abundance; the number of unique OTUs in each sample was represented by Observed Species and Shannon index. Based on these three metrics, Rarefaction curves were generated. The beta-diversity indexes, weighted and unweighted unifrac, were generated with QIIME. The Student’s t-tests was used to calculate the p value between the groups at the genus level, and p < 0.05 (*) was considered as statistically significant.

A random forest algorithm was used to predict the classification of samples for further exploring the relationship between fecal microbiota and CA. Random forest regression was performed with 1000 regression trees based on 5-fold cross-validation, 80% of the samples were randomly selected for model training and the remaining 20% were used for validation. The predicted result is displayed by the roc curve using R package: “proc”. In the random forest prediction classification algorithm, the contribution of different genera can be known.

### The 16S rRNA Gene-Based PCR Assay Specific for Faecalibacterium genus

The extracted genomic DNA was subjected to a PCR assay for detection of Faecalibacterium and all bacteria. Each reaction mixture comprised 2× SYBR Premix UrTaqTM II (Without Rox), 0.2 μM concentrations of the 16S rRNA gene specific primers: for Bacterial Universal primers, Forward: 5’-CGGTGAATACGTTCCCGG-3’, Reverse: 5’-TACGGCTACCTTGTTACGACTT-3’; for Faecalibacterium genus, Forward: 5’-GATGGCCTCGCGTCCGATTAG-3’, Reverse: 5’-CCGAAGACCTTCTTCCTCC-3’. and 20 ng of genomic DNA in a final volume of 20 µL. LightCycler® 480 II Instrument (Roche, Germany) thermal cycler used the following program: 95°C for 2 minutes, followed by 40 cycles of 10 s at 95°C, 30 s at 60°C, and 20 s at 70°C.

### Metabolites analysis with LC-MS

The samples were mixed with methanol, and were subjected to ultrasonic extraction in ice water for 10 min and centrifuged at 12,000 rpm at 4°C for 15 minutes, then the supernatant was harvested. Untargeted metabolite screening was performed on Q Exactive Orbitrap mass spectrometer (Thermo Scientific, Waltham, MA, USA) after calibration following the manufacturer’s guidelines. Full ms ddMS2 method was used to select QC samples, and multiple scanning segments were set to obtain more secondary data for identification. The samples were operated in both the positive and negative ion modes. Standardized samples were prepared for quality control.

Mass spectrometry data were converted to the mzML format using MS converter software. Then, XCMS (version 1.50) software was used to process data, including a peak search and peak alignment. Finally, OSI-SMMS software (version 2.0.0, Dalian ChemDataSolution Information Technology Co. Ltd) was used for substance identification. The positive and negative ion data were first calculated as the relative abundance, and those with an ms. score >0.5 were retained, combined and finally used for data analysis. The different metabolites were analyzed by Wilcoxon test analysis with R studio software (version 3.6.2) (p < 0.05). Pathway enrichment analysis of the differential metabolites was performed with the KEGG database (https://www.metaboanalyst.ca/home.xhtml). Pathways were considered significantly altered when p < 0.05. A network between different genera and all or different metabolites was generated by R studio software and visualized by Cytoscape (version: 3.6.1) (correlation>|0.6|, p<0.05). The Pearson correlation coefficient was calculated by psych (1.9.12.31), and the network was visualized by Cytoscape (version: 3.6.1) (p value < 0.05).

### Statistical data analysis

The data were represented as mean±standard deviation (SD), and each experiment had triplicate data. The Student’s t-tests was used to calculate the p value between the groups, and p < 0.05 (*) was considered as statistically significant.

## Results

### 1. Baseline characteristics of the recruited participants

To investigate the gut microbiota in cerebral aneurysm patients, a total of 140 patients and 40 healthy controls were recruited. Of those patients, 85 saccular aneurysm (A) and 23 dissecting aneurysms (D) were included in this study. All individuals recruited in the CA patient group were identified by Digital Subtraction Angiography (DSA) (**Supplemental Figure 1**). The characteristics of the patients and aneurysms are summarized in **Table 1**, including age, sex, surgery, stent, complication, Hemorrhage, Thrombus, Days, Aneurysm Size as well as post-surgery prognosis mRS1 (Period of hospitalization) and mRS2 (5 months after surgery). The differences in blood routine parameters between aneurysm patients and healthy controls are also shown in **Figure 1**. There were divergences in 5-nucleotidase, adenosine deaminase, chlorine, D-dimer, erythrocyte, mean erythrocyte width, hematocrit and prothrombin standardization ratio existed in the blood of patients with A or D to that of healthy people. The above variations represented the differences in Cardiac, liver, coagulation, erythrocyte and Electrolyte-related functions, respectively (**Figure 1**).

**Figure 1.**
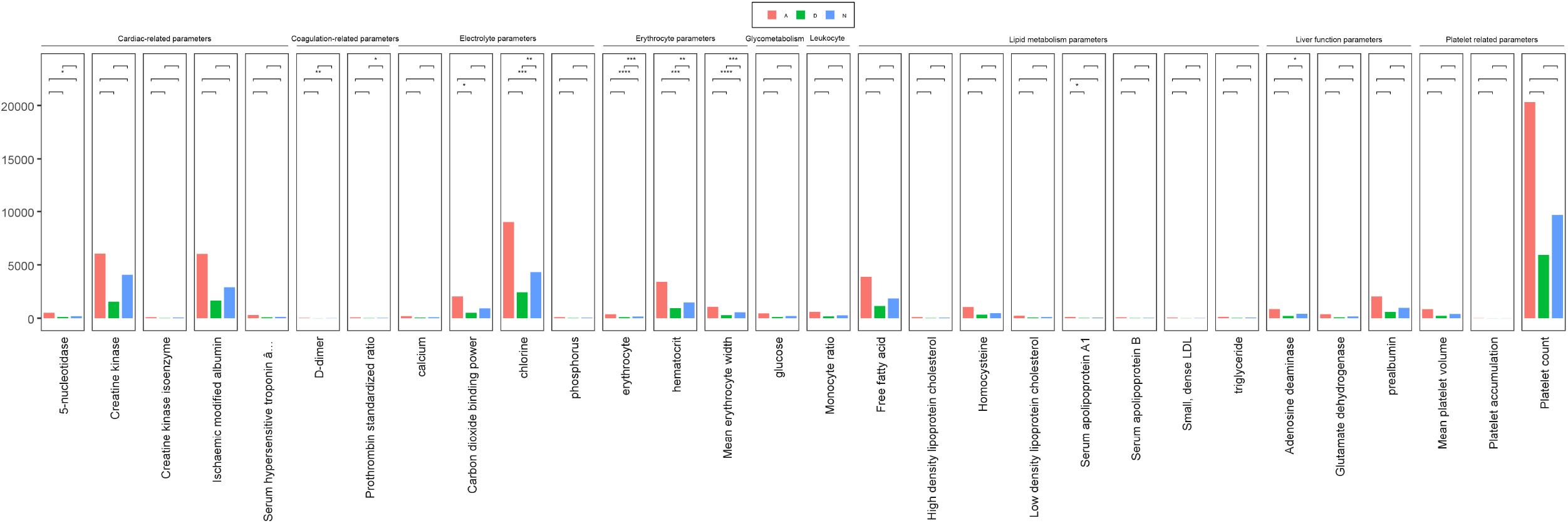
The Differences in blood routine parameters between patients with cerebral aneurysm subtype saccular aneurysm (A) and dissection aneurysm (D) and healthy controls (N). *, <0.05; **, <0.01; ***, <0.001; ****, <0.0001.

### Shifting in intestinal microbiota after the occurrence of CA

To study the variations in fecal bacteria after the occurrence of cerebral aneurysm, stools from the recruited patients of cerebral aneurysm and healthy people were collected and 16S rRNA sequencing and metabolomics analysis were performed on all samples, detailed information is introduced in the Material and Methods section. The rarefaction curve of indicated that the observed features of all samples tended to saturate as the sequence number increased (**Supplemental Figure 2**), indicating that the OTUs in the data cover most of the reads in the samples.

Succeedingly, the t-test of the patients of cerebral aneurysm showed that α-diversity indices of saccular aneurysm and dissecting aneurysms patients were higher than those of healthy people (p value<0.05) (**Fig. 2A**), including the Chao1estimated richness, observed features, as well as Shannon’s and Simpson’s diversities, which confirmed the altered within-sample microbial diversities between patients and healthy people. Similarly, a weighted_unifrac_t-test of faecal microbiota β-diversity demonstrated a significant variation between the patients and healthy control groups (**Fig. 2B**, p<0.01). The PCoA diagram verified the results of the weighted_unifrac_t-test of β-diversity, and the p-values obtained by ADONIS (permutational MANOVA) analysis of the bacterial variations were <0.001 (**Supplemental Table 1**, Pr value), suggesting that change in the between-sample microbial diversities of the intestinal microbiota was associated with the occurrence of cerebral aneurysm. It is noteworthy that the α- and β-diversity indexes showed no significant differences between the patients of saccular aneurysm and dissecting aneurysms, indicating the consistency in the microbial richness, evenness and relative abundance of the gut microbiota of cerebral aneurysm patients (**Supplemental Figure 3**). The above mentioned results suggested that both within-sample and between-sample microbial diversities of the microbiota were significantly shifted when cerebral aneurysm occurred.

**Figure 2.**
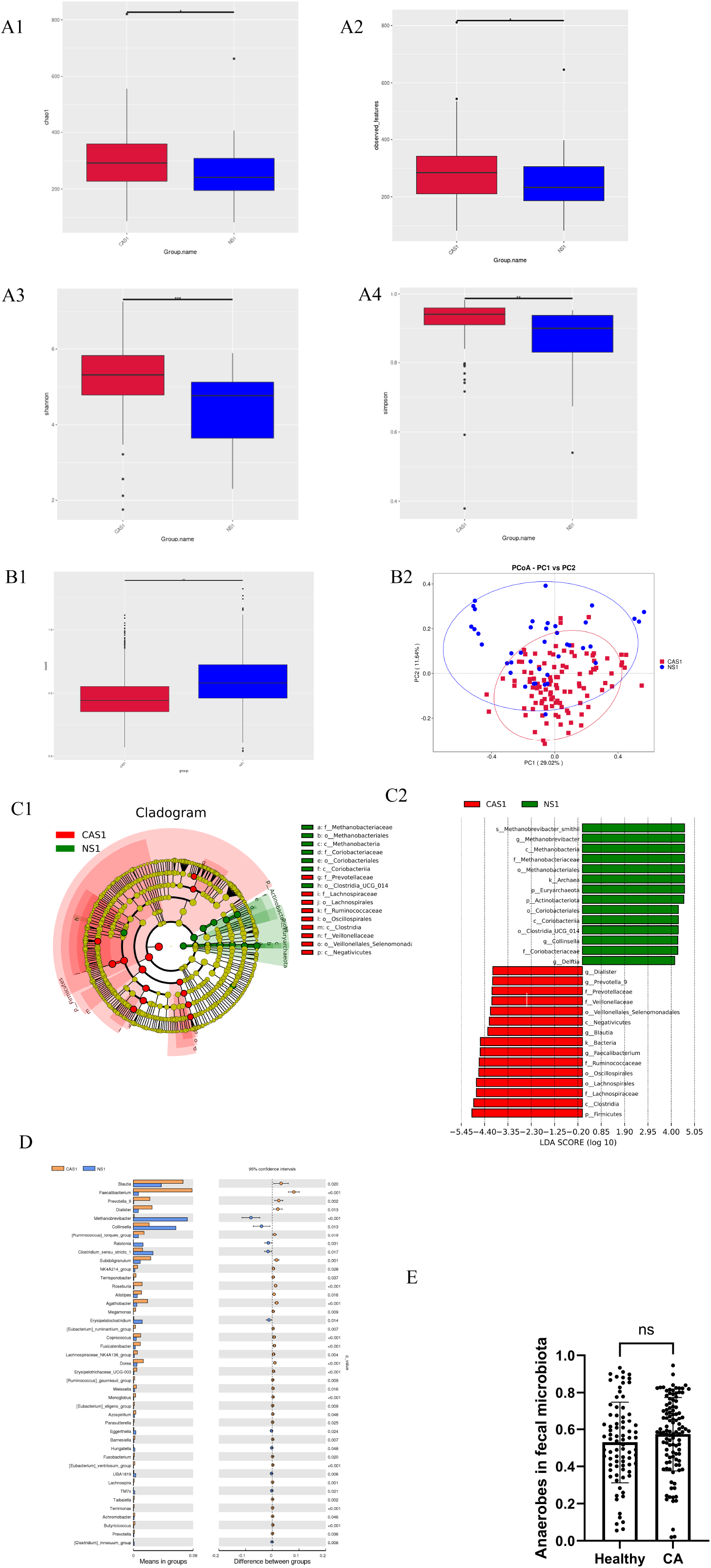
Variations of intestinal microbiota after CA. (A) Changes in microbial α-diversity, including richness estimated by chao1 estimated richness, observed features, Shannon’s and Simpson’s diversity; (B) Differences in β-diversity, illustrated with weighted_unifrac_t-test and PCoA diagrams; (C) LEfSe analysis of intestinal microbiota; (D) Bacteria with significant differences and relatively high abundance; (E) obligate anaerobe contents in the the CA and healthy control groups. *, <0.05; **, <0.01; ***, <0.001; ****, <0.0001.

To clarify the effects of CA on fecal microbiota, we then compared the structure of fecal microbiome at various taxonomic levels. The dominant phyla of healthy people and cerebral aneurysm groups were the Firmicutes and Bacteroides, these two phyla accounted for 43.2±23.6% and 14.1±20.5% of the microbiota in the control group, respectively; while in the patients group, the contents changed to be 61.2±20.6% and 14.4±15.4%, respectively (**Supplemental Figure 4A**). At the genus level, Faecalibacterium and Escherichia-Shigella were the two most abundant genera in the patients group, while the abundances of Bacteroides and Bifidobacterium dominated in the control group (**Supplemental Figure 4B**). LEfSe analysis revealed that Genus Blautia (k_Bacteria;p_Firmicutes;c_Clostridia;o_Clostridiales;f_Lachnospiraceae;g_Blautia), Dialister (k_Bacteria; p_Firmicutes; c_Clostridia; o_Clostridiales; f_Veillonellaceae; g_Dialister), Faecalibacterium (k_Bacteria; p_Firmicutes; c_Clostridia; o_Clostridiales; f_Ruminococcaceae; g_Faecalibacterium), Prevotella (k_Bacteria; p_Bacteroidetes; c_Bacteroidia; o_Bacteroidales; f_Prevotellaceae; g_Prevotella) as well as some of their parent taxonomic units were significantly overrepresented in the fecal microbiota of cerebral aneurysm patients (**Fig. 2C**), while Methanobrevibacter (k_Archaea; p_Euryarchaeota; c_Methanobacteria; o_Methanobacteriales; f_Methanobacteriaceae; g_Methanobrevibacter) and Collinsella (k_Bacteria; p_Firmicutes; c_Clostridia; o_Coriobacteriales; f_Coriobacteriaceae; g_Collinsella) related taxonomic units were overrepresented in the controls. The Lefse results were verified by t-test analysis, and their differential bacteria with high abundances were consistent (**Fig. 2D**). Interestingly, compared to the control group, where no sample had Faecalibacterium content greater than 10%, 32.9% (27/85) of saccular aneurysm and 30.4 (7/23) of dissecting aneurysms exceeded this value. However, the contrast between saccular aneurysm and dissecting aneurysms group was minimal, significant differences only existed in the contents of f__Veillonellaceae and O_Lactobacillales (**Supplemental Figure 5**). The above results showed that the intestinal microbiota changed significantly after CA. Although the abundance of some genera obligate anaerobic bacteria decreased in the patients group, such as g__Methanobrevibacter and g__Collinsella, the content of some other anaerobic genera increased, such as g_Blautia, g_Dialister, g_Faecalibacterium and g_Prevotella (**Figure 2D**), therefore, the proportion of obligate and facultative anaerobic bacteria in the microbiota might not change in general; suggesting the intestinal microenvironment of CA patients may have minimal changes in oxygen content. This hypothesis is further confirmed by the resemble contents of obligate anaerobes between the two groups of samples (**Figure 2E**).

### Effects of CA on fecal metabolites

To interrogate the correlation of metabolites and cerebral aneurysm, LC-MS system was employed to study variations in fecal metabolites between the patients and healthy controls. After confirming the data quality control, 941 anion and 1856 cationic metabolites present in the stool of healthy and aneurysmal patients were identified, **Supplemental Figure 6** illustrated the broad categories to which they belong. Partial Least Squares Discriminant Analysis (PLS-DA) was used to analyze the metabolites with differences between the patients and the control groups. Significant differences were found in the PLS-DA of positive ion to that of negative ion (**Figure 3A**), most of the samples were separated, indicating that differences were existed among the contents of metabolites between the two groups. Based on the Variable Importance in the Projection (VIP) value, some metabolites with possibly greater contributions to cerebral aneurysms have been found, such as negative ion sedoheptulose 7-phosphate (S-7-P), N4-(5-chloro-4-methoxy-3-thienyl)-2,6-dimethylmorpholine-4-carboxamide, sedoheptulose 1,7-bisphosphate (S-1,7-BP) and geranyl pp, as well as positive ion N-{6-[(5-chloro-3-pyridyl)oxy]-3-pyridyl}-N’-methylurea, pyritinol, N-Methyltryptamine and cinchophen (**Figure 3B and Supplemental Figure 7**). The differential metabolites were then imported into Kyoto Encyclopedia of Genes and Genomes (www.kegg.jp) to perform metabolic pathway matching analysis and to identify the cerebral aneurysms related metabolic pathways. **Figure 3C** summarized the enriched metabolic pathways, Carbon metabolism and Central carbon metabolism in cancer were the signaling pathways with the lowest p-value in anionic and cationic metabolites, respectively. The above metabolite enrichment results highlight the role of carbohydrate metabolism in cerebral aneurysm.

**Figure 3.**
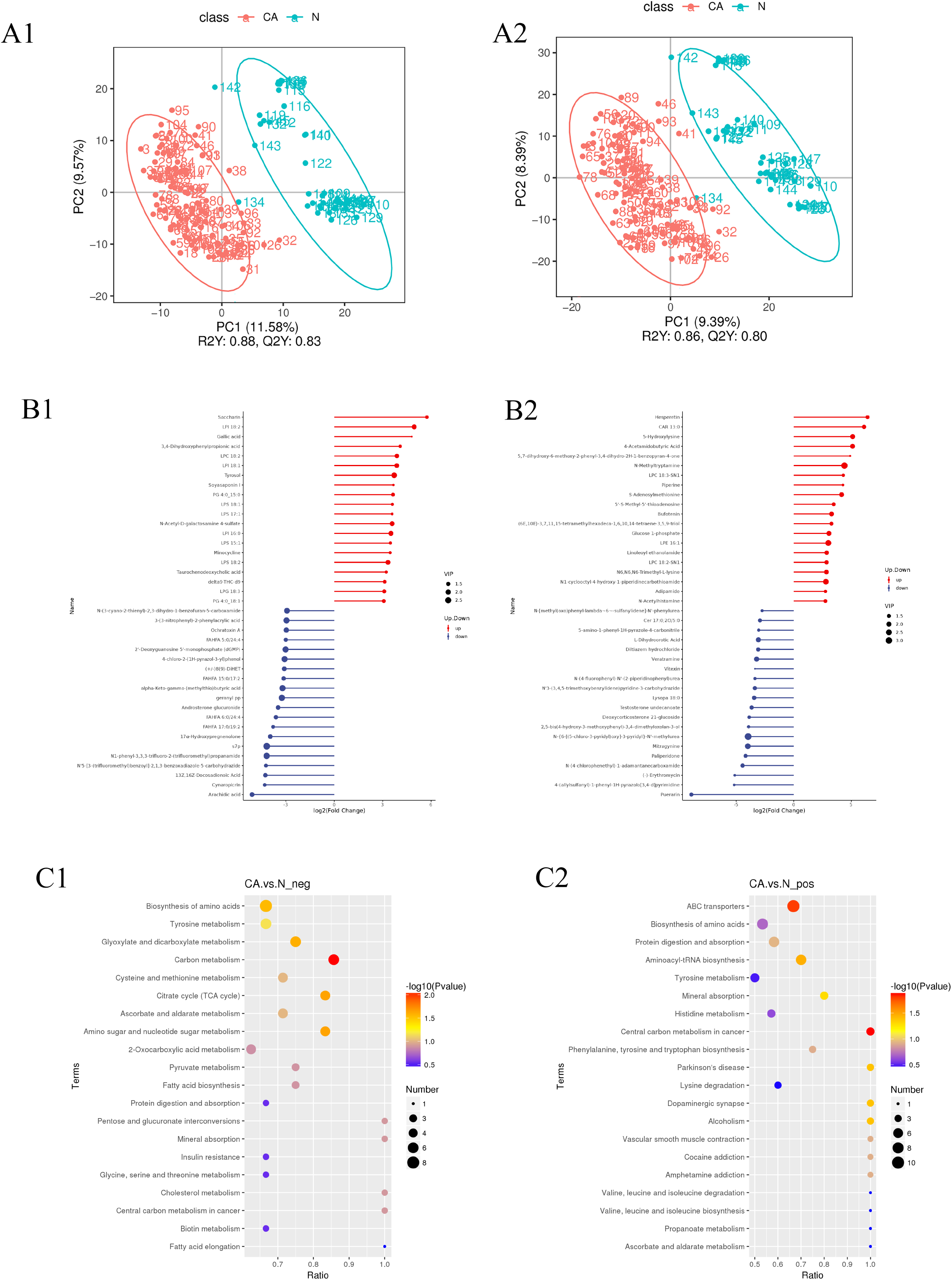
Changed fecal metabolites in CA patients. (A) PLS-DA divergences in anion and cationic metabolites; (B) Variable Importance in the Projection (VIP) values; (C) Enriched metabolic pathways in anion and cationic metabolites.

### Association between the differential microbiota, metabolites and blood parameters

To investigate the interrelationships of gut microbiota, metabolites and blood routine parameters, the Spearman’s correlation analysis was utilized to analyze the association them in the samples of healthy people and patients. **Figure 4A** demonstrated the significant association between the differential genus and the top discriminative metabolites with higher VIP values. After filter out the weak associations with |Spearman correlation coefficient| <0.4 and Q-value>0.01, genus Faecalibacterium and Roseburia stand out and they were interlinked (**Supplemental File 1**, correlation and p values). In addition to both being negatively related to metabolites sedoheptulose-7-phosphate (s7p), N-{6-[(5-chloro-3-pyridyl) oxy]-3-pyridyl}-N’-methylurea, N4-(5-chloro-4-methoxy-3-thienyl)-2,6-dimethylmorpholine-4-carboxamide, sedoheptulose-1,7-bisphosphate, geranyl-pp, 3-(4-chlorophenyl)-5-methyl-2,5-dihydro-1, 2, 4-oxadiazole, 4-chloro-2-(1H-pyrazol-3-yl) phenol, N-(1,2,3,4-tetrahydro-1-naphthalenyl) benzenesulfonamide and N-(3-cyano-2-thienyl)-2,3-dihydro-1-benzofuran-5-carboxamide, 4-methyl-2-Oxopentanoic Acid, N-methyltryptamine, Faecalibacterium was individually related to N1-phenyl-3,3,3-trifluoro-2-(trifluoromethyl) propanamide, 2’-Deoxyguanosine 5’-monophosphate (dGMP), lapachol, oxazepam-d5 and tyrosol, while Roseburia was correlated with 3-(3-nitrophenyl)-2-phenylacrylic acid, 5-(2-pyridinyl)-N-[2-(trifluorome-thyl)phenyl]-2-thiophenesulfonamide, and 5-[4-(phenylsulfonyl)phenyl]-1H-pyrazole. Moreover, Butyricicoccus was also found to related to tyrosol and 3,5-dimethyl-4H-pyrazol-4-one 4-(3,5-dichlorophenyl)hydrazine with relatively high correlation and low p values. These results indicate that the above said bacteria likely play critical regulatory roles in the carbohydrate metabolism during the occurrence of cerebral aneurysm.

**Figure 4.**
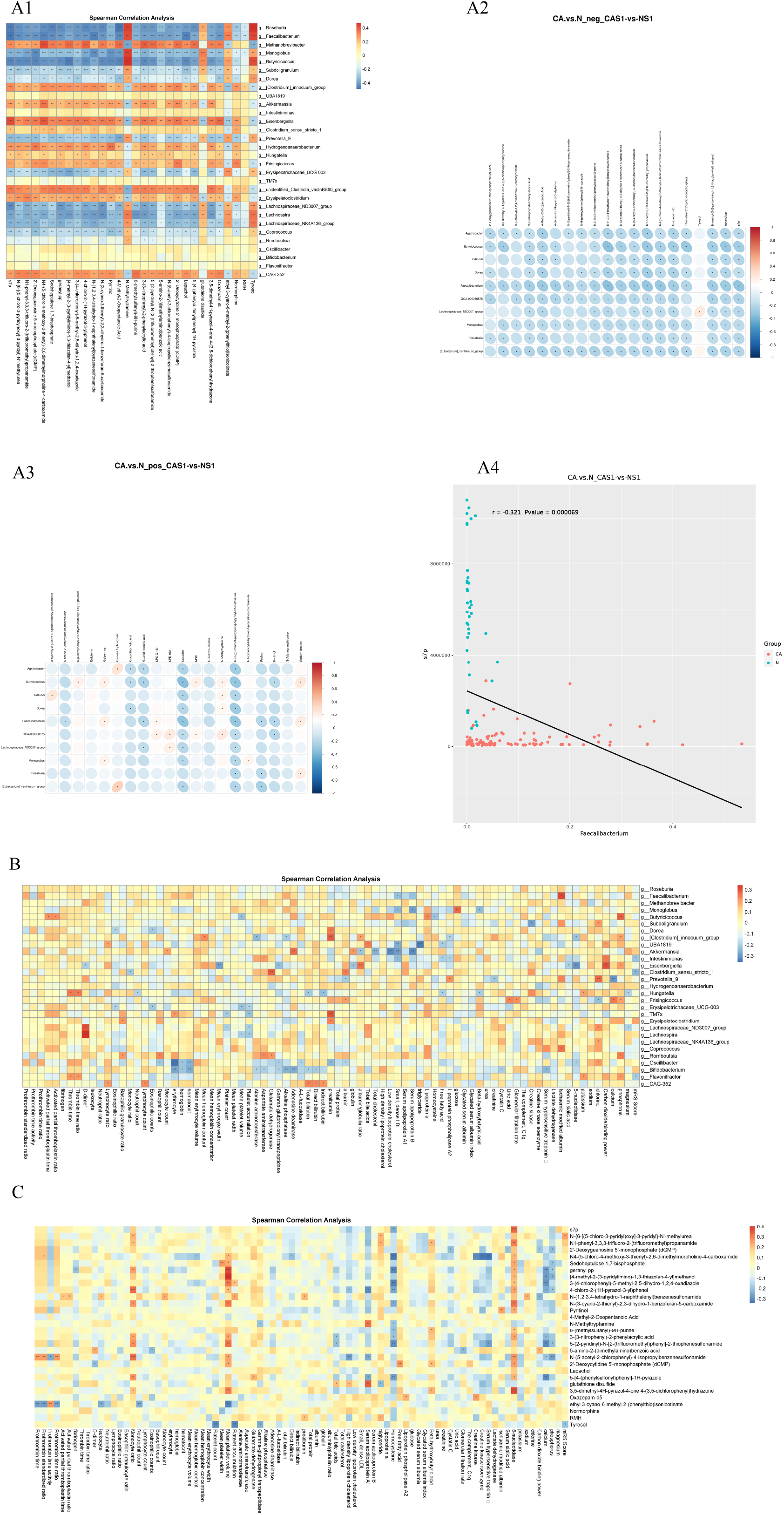
Associations between differential microbiomes, metabolites, and blood parameters. (A) Significant interactions between altered bacterial genera and metabolites with higher VIP values; (B) Correlation of fecal bacteria and metabolites with blood parameters; (C) Association of metabolites with blood parameters. *, <0.05; **, <0.01; ***, <0.001; ****, <0.0001.

Then the correlation of fecal bacteria and metabolites with blood routine parameters was investigated. The data demonstrated that Bifidobacterium had the most associations with blood parameters, including the amounts of erythrocyte, hemoglobin, hematocrit, Alanine aminotransferase, Aspartate aminotransferase, Gamma-glutpropionyl transpeptidase, Alkaline phosphatase, Adenosine deaminase, Total bilirubin, Direct bilirubin, Cystatin C and Serum hypersensitive troponin. Oscillibacter, which was second to Bifidobacterium, and the blood parameters associated with it were Basophil count, erythrocyte, hemoglobin, hematocrit, Gamma-glutpropionyl transpeptidase, A-L-fucosidase, prealbumin, albumin, 5-nucleotidase and chlorine (**Figure 4B**). It is noteworthy that Faecalibacterium associates with Small dense LDL and Ischaemic modified albumin, which have been shown to be associated with atherosclerosis or ischemic stroke in other diseases. Reversely, chlorine and phosphorus had the most correlation generas among the blood parameters, chlorine associated with genus Subdoligranulum, Prevotella, Frisingicoccus, Lachnospiraceae and Oscillibacter, while phosphorus associated with genus Butyricicoccus, Clostridium, Eisenbergiella, Frisingicoccus, Romboutsia and Flavonifractor.

For the interaction of metabolites with blood parameters, N4-(5-chloro-4-methoxy-3-thienyl)-2,6-dimethylmorpholine-4-carboxamide and N-(5-acetyl-2-chlorophenyl)-4-isopropylbenzenesulfonamide had the most associations with blood parameters, its abundance associated with the amount of Prothrombin standardized ratio, Mean hemoglobin content, Mean platelet volume, Direct bilirubin, Homocysteine, Creatine kinase, Creatine kinase isoenzyme, Serum hypersensitive troponin, 5-nucleotidase and the amount of Prothrombin time, Prothrombin standardized ratio, Prothrombin time activity, Prothrombin time ratio, Monocyte ratio, Mean platelet volume, High density lipoprotein cholesterol, Serum apolipoprotein A1, 5-nucleotidase, calcium, phosphorus, respectively (**Figure 4C**). It is noteworthy that s7p, the molecule associated to genus Faecalibacterium and Roseburia, correlated with Homocysteine and 5-nucleotidase. Homocysteine molecule is an independent risk factors for stroke, while 5-nucleotidase is an indicators of liver function. The above correlation results indicated that some intestinal bacteria and metabolites are related to host blood parameters and may have crucial physiological functions in CA occurrence.

### Using differential bacteria and metabolites to distinguish patients of CA from healthy people

To explore the feasibility of using differential bacteria and metabolites to assess who were at a higher risk of cerebral aneurysm, a random forest classifier model was applied for group prediction. The differential fecal bacteria, metabolites and blood parameters were first evaluated by random forest models, and the results exhibited that fecal bacterial genus Roseburia (ASV23), Faecalibacterium (ASV5), fecal metabolite s7p (META2781) as well as blood routine parameter erythrocyte (clinic9) had the greatest influences on the accuracy of certain cerebral aneurysm predictive model (**Figure 5A**). Those factors with a relatively high Mean Decrease Accuracy, were then combined randomly to construct predictive models for predicting cerebral aneurysm, and the results showed that the ROC of model built by the combination of g__Faecalibacterium, s7p and Mean erythrocyte width obtained the highest AUC up to 99.8% (**Figure 5B**). However, analysis of this predictive model showed that Pr (>|z|) values of g__Faecalibacterium, s7p and Mean erythrocyte width were all >0.05 (**Supplemental Table 2**). If only g__Faecalibacterium and s7p were used to construct the predictive model, the AUC in ROC was 99.7% (**Figure 5C**). Meanwhile, Pr (>|z|) of g__Faecalibacterium and s7p were both <0.05, which are 0.020 and 0.0086, respectively (**Supplemental Table 3**). Moreover, the predictive model built with only g__Faecalibacterium obtained an accuracy of 82.8% and its Pr (>|z|) value is 0.001 (**Figure 5D and Supplemental Table 4**). Given that Faecalibacterium and s7p have the potential as screening biomarkers for CA and no suitable antibodies have been found currently to detect s7p, only the abnormal enrichment of Faecalibacterium was verified by Real-time PCR with 12 newly collected samples. The results showed that the detection accuracy was 91.7% (11/12) (**Figure 5E**). The above results exhibited the role of intestinal bacterium Faecalibacterium and metabolites s-7-p as potential biomarkers in CA prediction.

**Figure 5.**
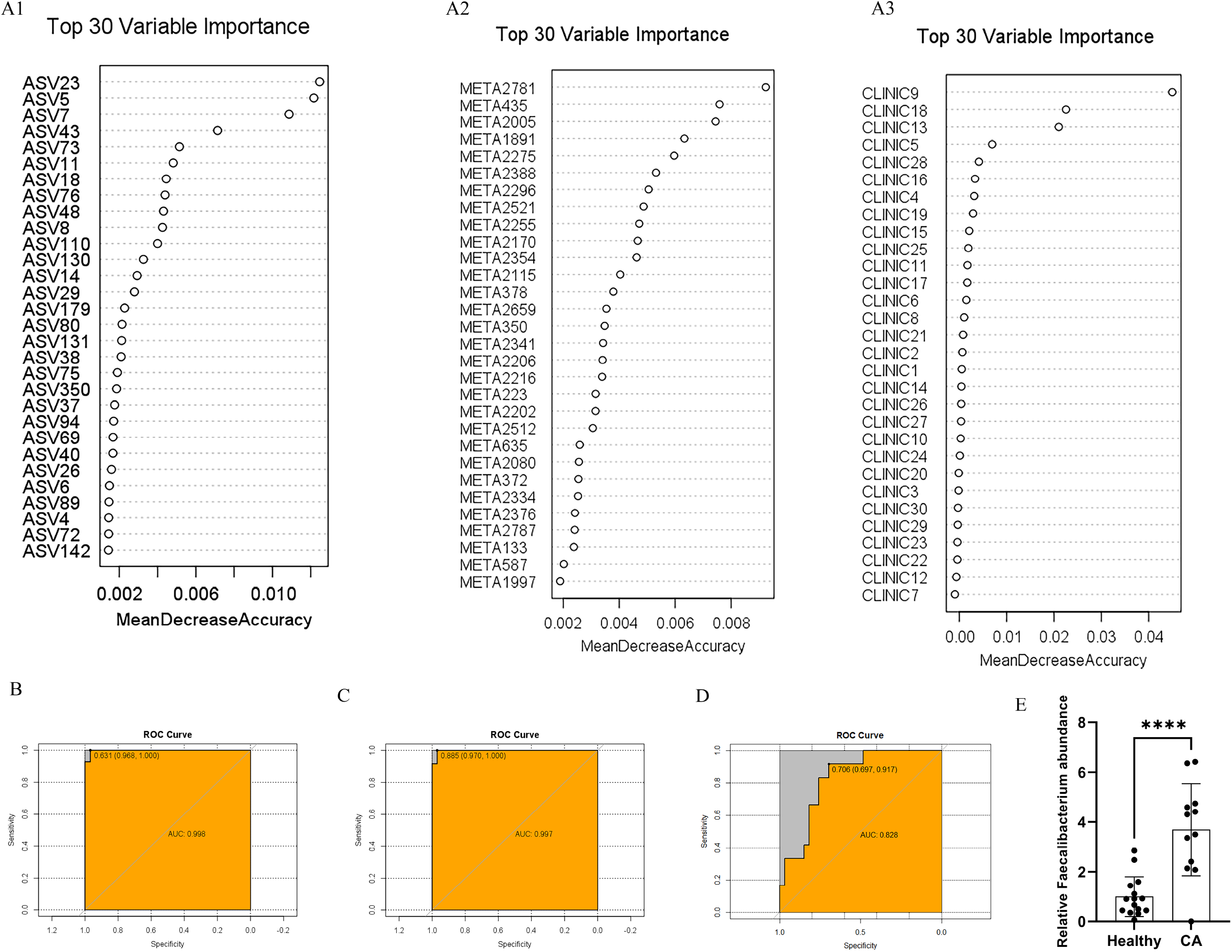
Bacteria and metabolites could be used to distinguish patients with CA from healthy individuals. (A) Screening and evaluation of the differential fecal bacteria, metabolites and blood parameters with random forest models; (B) The ROC curve of a predictive model constructed with the combination of g__Faecalibacterium, s7p and average red cell width; (C) The ROC curve of a predictive model constructed with faecalibacterium and s7p; (4) The ROC curve of a predictive model constructed with g__Faecalibacterium only; (E) Quantitative real-time PCR was performed to test the 12 newly collected samples for verifying the content of Faecalibacterium. *, <0.05; **, <0.01; ***, <0.001; ****, <0.0001.

### The value of Intestinal microbiota, metabolites and blood parameters in predicting CA prognosis

Next, we evaluated the feasibility of microbiota, metabolites, and blood parameters as potential biomarkers for predicting prognosis of cerebral aneurysm. Univariate and multifactorial Cox regression analysis were performed to estimate the correlation of post-surgery quality of life with various intestinal microbiota, metabolites and blood parameters. The results showed that only metabolite tyrosol and blood parameter mean platelet volume, total protein, albumin, albumin/globulin ratio and high density lipoprotein cholesterol had significant correlation with prognosis within 95% confidence interval (**Table 2**, p<0.05). Interestingly, although the correlation of Faecalibacterium or s-7-p to surgical outcomes was not significant under this limited sample size, the post-surgery prognostic related metabolite tyrosol was associated with Faecalibacterium directly (Figure 4A), while blood parameters mean platelet volume and high density lipoprotein cholesterol were correlated with metabolite geranyl pp, [4-methyl-2-(3-pyridylimino)-1,3-thiazolan-4-yl]methanol and N-(5-acetyl-2-chlorophenyl)-4-isopropylbenzenesulfonamide, the latter two metabolites were significantly associated with Faecalibacterium or s-7-p (**Figure 4A, C and Figure 6**). Conclusively, the results highlight the significance of altered intestinal bacteria and metabolites Faecalibacterium or s-7-p as potential biomarkers in CA prediction and post-surgery prognosis.

**Figure 6.**
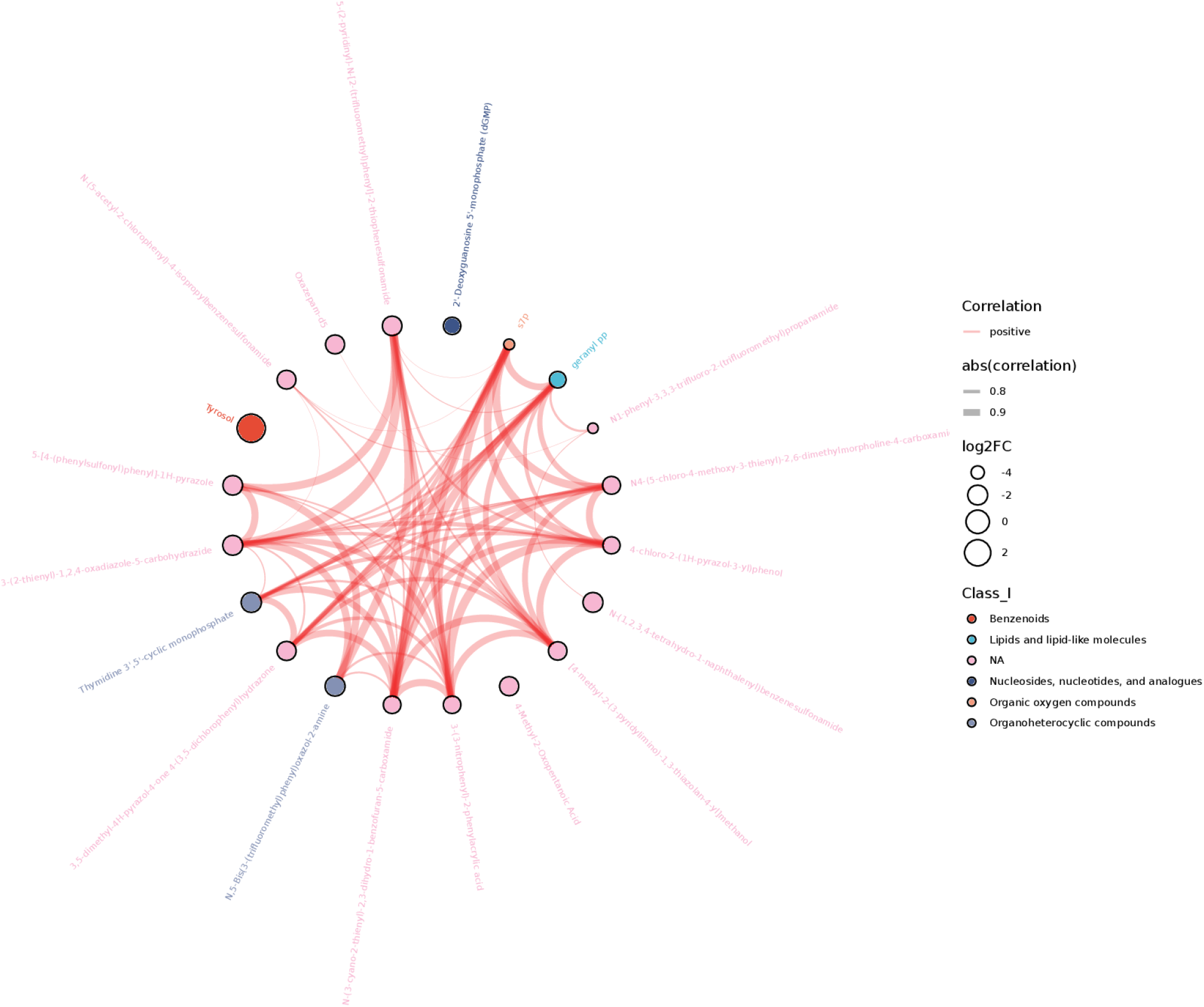
The metabolites that significantly associated with s-7-p.

## Discussion

An aneurysm is a permanent swelling occurs in the arteries of the cerebral, the aorta, and the major arteries that pump blood from the heart to the rest of the body. Unruptured intracranial aneurysms (UIAs) have a prevalence of ≈3%[1]. Once rupture, ≈50% of the patients die, and 50% of the surviving patients suffer from severe complications[17]. According to previous studies, a variety of factors affected the incidence of CA formation, including: genetic factors, arteriosclerosis, infection and trauma, etc. There is growing evidence that the gut microbiome affects host metabolism and immune homeostasis and may be a key risk factor for cardiovascular disease[18]. However, there is no direct causal relationship reported between microbial variation and the formation of CA. With the increasing popularity of MRI and CT examination after dizziness and other symptoms in the elderly, more and more CAs have been found. These suspected aneurysms require further examination, such as MRA, CTA, or DSA, to verify the diagnosis and exploring details of disease such as location, size, and morphology. Often in many cases, aneurysm monitoring is also required to evaluate treatment response and effect. However, previous CA detection techniques, especially MRA and CTA, have inadequate capability to detect aneurysms less than 4mm in diameter, with high false-positive and false-negative rates. Also, these examinations are often expensive and not applicable if the patient is allergic to the contrast agent. In contrast, the changes of human microbiota are closely related to the pathophysiological state of the body [13], therefore they are highly responsive to changes in the body. Microbial variations can produce detectable changes earlier than the appearance of disease symptoms, which is a sensitive biomarker for disease risk prediction. Exploring the relevance of the microbiota to disease can also help in the search for clues to disease mechanisms [14]. In the present investigation, we analyzed fecal microbes and their metabolites from CA patients and healthy controls and explored their correlation with blood parameters. The results indicated significant differences between the healthy control group and the CA group, not only in the gut microbiota, its metabolites and blood composition, but also in the interactions between them. After random forest model analysis and verification with QPCR, Faecalibacterium and s-7-p can predict aneurysms with an accuracy of 99.7%. Although Faecalibacterium and s-7-p did not achieve the expected significance in predicting post-surgery prognosis, they were associated with those metabolites that achieved significance. It is believed that Faecalibacterium and s-7-p even their related enzymes will highly likely be associated with post-surgery prognosis with the expansion of the sample size, which makes the combination of bacteria and metabolites as promising non-invasive, inexpensive and sensitive biomarkers for the early detection and post-surgery prognostic of CA.

Treatment options for CA is either follow-up observation or surgical treatment. In general, aggressive surgical intervention is preferred for those patients with saccular aneurysms or dissecting aneurysms ≥5mm in diameter. The commonly used surgical procedures for CA include craniotomy clipping and interventional surgery, such as coil embolization and shunt implantation. However, it has also been reported that some CAs with a diameter of less than 5mm also have the risk and possibility of rupture and bleeding. The incidence of complications in surgical treatment of aneurysms is about 3-5%, such as intracranial hemorrhage, cerebral vasospasm, and cerebral infarction. Therefore, it is still imperative to further study the internal mechanism of CA formation and rupture. This study demonstrated us that gut bacteria and their metabolites are associated with blood parameters, especially those cardiac and vascular-related markers. In the future, the relationship between these factors and inflammation and atherosclerosis might be furtherly elucidated, and could provide us with a microbiome based mechanism for CA occurrence or recovery, even possibly develop a set of microbial therapeutic strategies as a clinical treatment option, such as probiotics usage and fecal bacteria transplantation etc.

The genus Faecalibacterium that identified divergently expressed in this research with predictive value is a beneficial butyrate-producing bacterium with anti-inflammatory properties that promotes intestinal homeostasis [19], it is characterized as a low-GC, gram-positive, non-spore-forming, strictly anaerobic, and non-motile firmicute [20]. Faecalibacterium is extremely sensitive to oxygen and is the most important butyrate producing bacteria in the human colon, butyrate is a key modulator of inflammation and colorectal cancer. In addition, it can also ferment glucose into acetate, D-lactate and formate [21, 22]. Although it has often been reported that pathological processes are significantly negatively correlated with the number of Faecalibacterium, it is also possible to show an elevation in certain circumstances to suppress inflammation. For example, Faecalibacterium level is high in the surviving colorectal cancer group, and it is negatively correlated with the expression of β-catenin, MMP-9 and NFκB [23]. Faecalibacterium is well adapted to the local intestinal microecosystem, where it may be cross-fed with other members of gut microbiota. However, the role of Faecalibacterium in CA remains poorly understood. In this study, Faecalibacterium was found not only associated with a variety of bacteria, metabolite, and blood factors in the microbiota, but also as a potential predictor of CA formation (Figure 3, 5 and Supplemental Figure 7). However, when studying its relationship with post-surgery prognosis, the results showed no significant correlation. This is probably due to insufficient sample size, and future clinical studies with larger sample size are needed to confirm the correlation between Faecalibacterium and prognosis. In the future, animal experiments will also be needed to clarify action mechanism of this bacterium according to its associated molecules and bacteria, and to pave the way for microbiota-based therapy.

Sedoheptulose-7-phosphate (s-7-p), an important intermediate in the non-oxidizing part of the pentose phosphate pathway, is a phosphorylated monosaccharide containing 7 carbon atoms and carbonyl functional groups. In addition to participating in intracellular primary metabolism, s-7-p is also involved in the natural synthesis of different C7N aminocyclitols by cyclase into different structural units. It was found that the cyclitols in acarbose, an α-glucosidase inhibitor for the treatment of type 2 diabetes, as well as Jinggangmycin A and its derivatives, are all formed from seven carbon sugars derived from s-7-p [24, 25]. Subsequent studies have also found that the cyclitols portion of the antibiotic pyralomicin 1a and the anti-tumor drug cetoniacytone A come from s-7-p [26, 27]. Moreover, s-7-p is revealed participating in the biosynthesis of outer membrane components of Gram-negative bacteria, such as lipopolysaccharide [28, 29]. However, the role of s-7-p in CA remains poorly understood. In our study, s-7-p and its related bacteria Faecalibacterium were not only identified as a potential predictor of CA, but also associated with c__blood parameters mean platelet volume and compound geranyl pp, [4-methyl-2-(3-pyridylimino)-1,3-thiazolan-4-yl]methanol and N-(5-acetyl-2-chlorophenyl)-4-isopropylbenzenesulfonamide, which have significances for post-surgery prognosis. More interestingly, s-7-p can also use certain enzymes to convert to some of the metabolites, which can provide clues for later tests that include the enzyme. In the future, its associated molecules and bacteria as well as action mechanism will be elucidated, the goal is to remove obstacles for microbiota-based detection and therapy.

## Conclusions

The current study presents variations in gut microbiota and its metabolites as well as blood routine parameters after CA, highlighting the significance of altered intestinal bacteria and metabolites as potential biomarkers in CA prediction and post-surgery prognosis.

## Data Availability

Availability of data and material http://www.ncbi.nlm.nih.gov/bioproject/1053898 Supplemental files: meta_intensity_class_neg and meta_intensity_class_pos

## Acknowledgements

The authors would like to thank Yang Shen for his assisting analysis of the data and for helpful discussions.

## Funding

This research was sponsored by “Design and development of a smart department management platform based on big data” (Shandong University No.6010122150).

## Declarations

### Ethics approval and consent to participate

This study was reviewed and approved by the Ethic Committee of School of Basic Medical Science, Shandong University (Jinan, Shandong Province, China), the approval number is ECSBMSSDU2023-1-82. All volunteers gave their written informed consent prior to their inclusion of the study. All methods were performed in accordance with the relevant guidelines and regulations.

## Competing interests

Authors declare that they have no competing interests.

## Authors’ contributions

J.D., G.L., D.W., P.Z, and P.Z. collected the samples; S.L. prepared the figures and wrote the main manuscript text; T.D. assembled the figures; L.D., Z.D. and S.L. designed the research. All authors have reviewed the final version of the manuscript and approved this submission.

## Availability of data and material

http://www.ncbi.nlm.nih.gov/bioproject/1053898

Supplemental files:

meta_intensity_class_neg and meta_intensity_class_pos

## Notes

### Competing Interest Statement

The authors have declared no competing interest.

### Author Declarations

This study was reviewed and approved by the Ethic Committee of School of Basic Medical Science, Shandong University (Jinan, Shandong Province, China), the approval number is ECSBMSSDU2023-1-82.

